# Lessons from the COVID-19 pandemic: People’s experiences and satisfaction with telehealth during the COVID-19 pandemic in Australia

**DOI:** 10.1101/2020.09.10.20192336

**Authors:** JMJ Isautier, T Copp, J Ayre, E Cvejic, G Meyerowitz-Katz, C Batcup, C Bonner, RH Dodd, B Nickel, K Pickles, S Cornell, T Dakin, K McCaffery

**Author notes:** **Correspondence:** Jennifer Isautier Rm 128A, Edward Ford Building A27 | The University of Sydney | NSW | 2006 | E.

## Abstract

**Objectives:** To determine how participants perceived telehealth consults in comparison to traditional in-person visits, and to investigate whether people believe that telehealth services would be useful beyond the pandemic.

**Design:** A national cross-sectional community survey.

**Participants:** Australian adults aged 18 years and over (n=1369).

**Main outcome measures:** Telehealth experiences.

**Results:** Of the 596 telehealth users, the majority of respondents (62%) rated their telehealth experience as “just as good” or “better” than a traditional in-person medical appointment. On average, respondents perceived that telehealth would be moderately to very useful for medical appointments after the COVID-19 pandemic is over (M=3.67 out of 5, SD=1.1). Being male (p=0.007), having a history of both depression and anxiety (p=0.037), or lower patient activation (individuals’ willingness to take on the role of managing their health/healthcare) (p=0.037) were associated with a poorer telehealth experience. Six overarching themes were identified from free-text responses of why telehealth experience was poorer than a traditional in-person medical appointment: communication is not as effective; limitations with technology; issues with obtaining prescriptions and pathology; reduced confidence in doctor; additional burden for complex care; and inability to be physically examined.

**Conclusions:** Telehealth appointments were reported to be comparable to traditional in-person medical appointments by most of our sample. Telehealth should continue to be offered as a mode of healthcare delivery while the pandemic continues and may be worthwhile beyond the pandemic.

The known
- The COVID-19 pandemic has resulted in an increase in telehealth services.

The new
- The majority of telehealth users (62%) perceived their experience to be just as good or better than traditional in-person medical care; and that telehealth would be at least somewhat useful beyond the pandemic.
- Having a history of both depression and anxiety was associated with a poorer telehealth experience and in-person visits were frequently preferred over telehealth visits for mental health appointments.

The implication
- Telehealth should continue to be offered while the COVID-19 pandemic continues and may be worthwhile beyond the pandemic, however, telehealth may be less effective for mental health services.

## Introduction

The coronavirus (COVID-19) outbreak was officially declared a pandemic by the World Health Organization on March 11 2020. To help minimize the spread of the virus, healthcare systems have rapidly adopted alternative models for healthcare delivery, including telehealth services (1). This type of healthcare delivery minimizes the spread of the virus by providing health care services without the need for face-to-face contact, reducing the risk of exposure to COVID-19 for both patients and clinicians.

In response to the COVID-19 pandemic, the Australian Government introduced a temporary telehealth scheme on March 30 2020 to enable subsidised access to healthcare services via telephone or videoconferencing (2). Prior to the pandemic, telehealth consultations were restricted to rural and remote communities. This new scheme allowed all medical appointments with a variety of health professionals to be conducted via telehealth, regardless of rurality. As a result of this scheme, telehealth consults accounted for 36% of all services provided in April 2020 compared to 1.3% before the pandemic (3). At the end of April 2020, a nationally representative survey by the Australian Bureau of Statistics (ABS) (N=1,022) reported that 1 in 6 people used a telehealth service (17%); women were almost twice as likely as men to use telehealth (22% vs 12%); and persons with a chronic/mental health condition were twice as likely to have used a Telehealth service as those without (25% vs 13%).

However, 1 in 10 people (11%) reported to have a GP or health professional appointment cancelled or postponed (4). Cancelling or postponing appointments is concerning because reduced healthcare utilization during pandemics has been associated with poorer health outcomes, for example during the Ebola virus outbreak and SARS epidemic (5, 6). The increased uptake of telehealth services and increased cancelling or postponing of medical appointments warrants further investigation to better understand people’s experiences and satisfaction with accessing telehealth services during the pandemic. This is particularly necessary given the long-term outlook of COVID-19 - while some health services have returned to normal, continuing outbreaks may deter patients from accessing in-person care for some time (7).

Despite the increase in telehealth, little is known about people’s experience of telehealth services compared to traditional-in person visits during the COVID-19 pandemic in Australia. We investigated the experiences related to telehealth in a sample of Australians during the COVID-19 pandemic. Our aims were to determine how participants perceived telehealth consults in comparison to traditional in-person visits, and to investigate whether people believe that telehealth services would be useful after the pandemic. Furthermore, we investigated the sociodemographic and health-related factors associated with negative telehealth experiences.

## Methods

### Recruitment

The data used in this study are from a prospective longitudinal national survey launched in April 2020 exploring variation in understanding, attitudes, and uptake of COVID-19 health advice during the 2020 pandemic (8). Here, we report on data from the survey wave conducted over a 1-week period in Australia from June 5 to June 12 2020, using the online platform Qualtrics. Participants were aged 18 years and over, could read and understand English and currently residing in Australia. Recruitment was via paid targeted advertisements on social media (Facebook and Instagram). More details on recruitment are given elsewhere (8). Participants were given the opportunity to enter into a prize draw for the chance to win one of ten $20 gift cards upon completion of the survey. This study was approved by The University of Sydney Human Research Ethics Committee (2020/212).

### Measures

Sociodemographic variables including age, gender and educational status were collected, as well as self-reported chronic diseases and overall health. We assessed health literacy using the Newest Vital Sign (9) and digital health literacy using the eHealth Literacy Scale (eHeals) (10). The Consumer Health Activation Index (CHAI) (11) was used to determine patient activation (individuals’ willingness to take on the role of managing their health and healthcare). Remoteness and socioeconomic status of place of residence were derived from participant postcode (12). Participants were asked to indicate whether they had used telehealth services, and if so, how telehealth services compared to traditional in-person visits, if they experienced any barriers to using telehealth, and whether they have cancelled or postponed an appointment with a health professional (Box 1).

##### Box 1: Survey Items and Scoring Scale on Telehealth

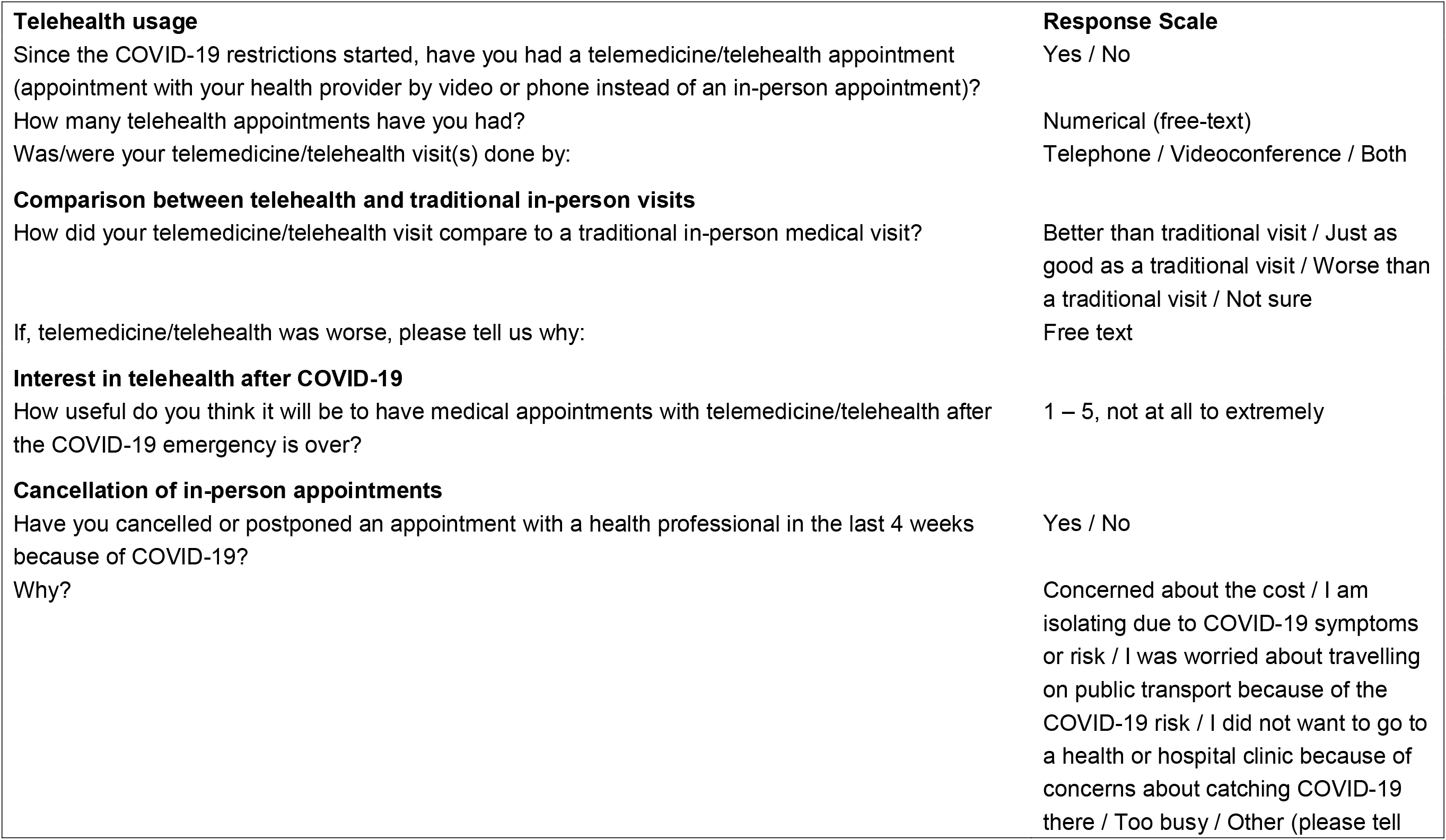
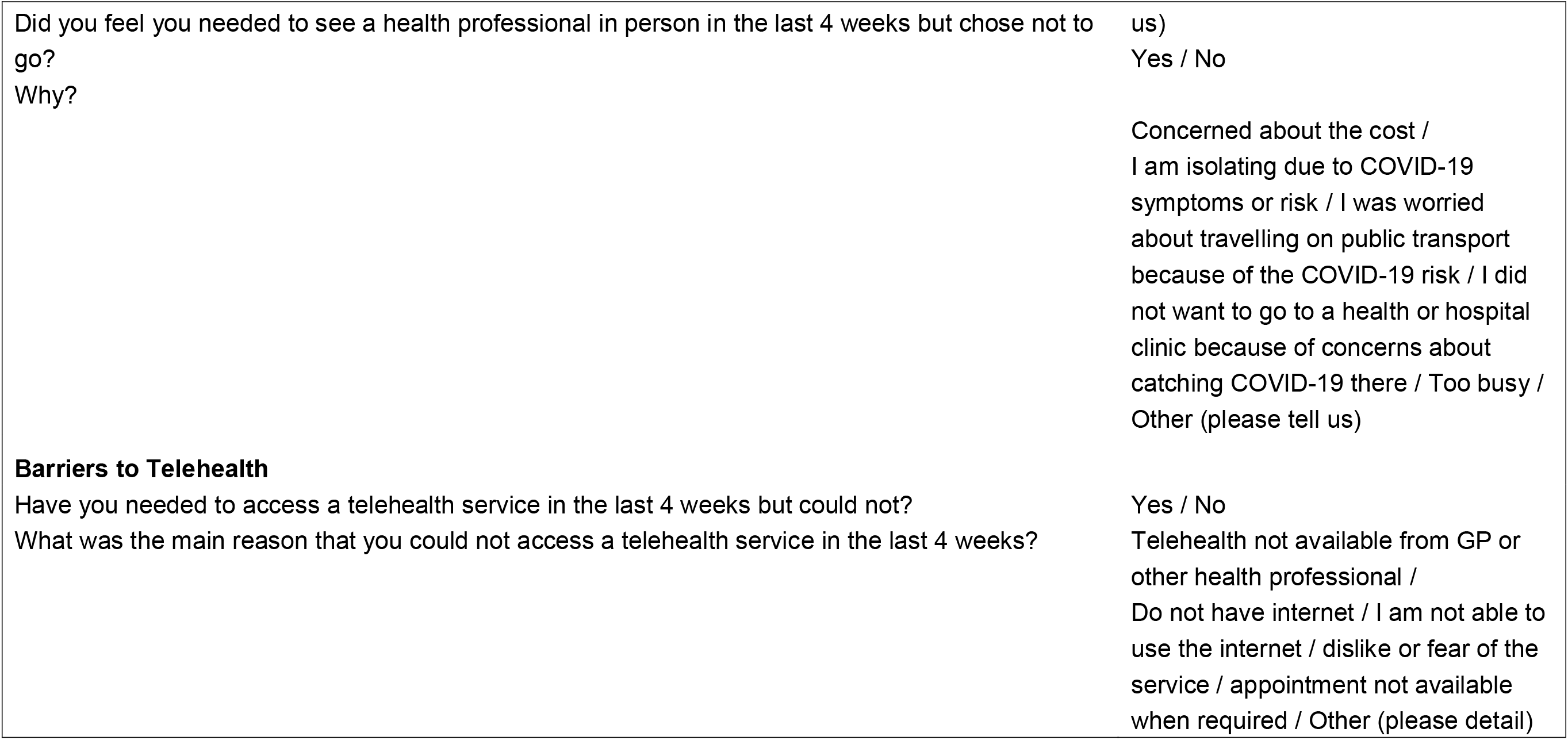

### Statistical analysis

Quantitative data were analysed using Stata/IC v16. IDescriptive statistics were calculated for sample characteristics and used to summarise the telehealth experiences of the sample since COVID-19 restrictions commenced. A generalised linear model utilising a modified Poisson approach (log link function with robust standard errors) was used to estimate adjusted relative risks (with 95% confidence intervals) of having a poorer telehealth experience compared to traditional in-person medical visits across various sociodemographic and health-related factors. An independent samples t-test was used to compare perceived usefulness of telehealth medical appointments once the COVID-19 emergency ends between those who rated their telehealth experience as worse, and those who reported the experience to be the same or better, than an in-person medical visit. Statistical significance for these exploratory analyses was set at p<0.05.

Qualitative data was analysed using content analysis (13) which combines both qualitative and quantitative methods, allowing both the frequency of categories to be reported and the content. JI and TC familiarised themselves with the content and generated a list of recurring themes; these were discussed and then checked by an additional researcher (JA). JI and TC then applied the final coding framework to all the data. The level of agreement was tested using Cohen’s kappa and indicated substantial agreement (κ=0.76) (14). Discrepancies were discussed until consensus was obtained. Descriptive statistics are provided to summarise the frequency of each code.

### Results

Of the 1,369 respondents who completed the June survey, 596 (45%) reported using telehealth services since the start of the pandemic. Those who used telehealth were slightly older, more likely to be female, had higher levels of education, greater prevalence of chronic health conditions (including mental health history), and poorer self-reported general health. Sample characteristics are summarised in Table 1.

**Table 1.** Descriptive characteristics of sample (N=1369) by use of telehealth during COVID-19 lockdown period. Data are shown as n (%) unless otherwise specified.

| Variable | Accessed telehealth services (n=596) | Did not access telehealth services (n=773) | Overall (N=1369) |
| --- | --- | --- | --- |
| Mean age (SD), years | 46.2 (16.1) | 43.6 (17.0) | 44.7 (16.7) |
| Age group |  |  |  |
| 18 to 25 years | 76 (12.8%) | 156 (20.2%) | 232 (16.9%) |
| 26 to 40 years | 166 (27.9%) | 206 (26.6%) | 372 (27.2%) |
| 41 to 55 years | 152 (25.5%) | 192 (24.8%) | 344 (25.1%) |
| 56 to 90 years | 202 (33.9%) | 219 (28.3%) | 421 (30.8%) |
| Gender |  |  |  |
| Male | 146 (24.5%) | 287 (37.1%) | 433 (31.6%) |
| Female | 433 (72.7%) | 478 (61.8%) | 911 (66.5%) |
| Other / prefer not to say | 17 (2.9%) | 8 (1.0%) | 25 (1.8%) |
| Highest level of educational attainment |  |  |  |
| High school or less | 68 (11.4%) | 130 (16.8%) | 198 (14.5%) |
| Certificate I-IV | 67 (11.2%) | 73 (9.4%) | 140 (10.2%) |
| University education | 461 (77.3%) | 570 (73.7%) | 1031 (75.3%) |
| Number of chronic health conditions<sup>^</sup> |  |  |  |
| None | 239 (40.1%) | 436 (56.4%) | 675 (49.3%) |
| One | 188 (31.5%) | 220 (28.5%) | 408 (29.8%) |
| Two or more | 169 (28.4%) | 117 (15.1%) | 286 (20.9%) |
| Mental health history |  |  |  |
| Depression | 278 (46.6%) | 193 (25.0%) | 471 (34.4%) |
| Anxiety | 302 (50.7%) | 232 (30.0%) | 534 (39.0%) |
| Self-reported General health |  |  |  |
| Poor | 37 (6.2%) | 9 (1.2%) | 46 (3.4%) |
| Fair | 111 (18.6%) | 76 (9.8%) | 187 (13.7%) |
| Good | 226 (37.9%) | 237 (30.7%) | 463 (33.8%) |
| Very Good | 172 (28.9%) | 321 (41.5%) | 493 (36.0%) |
| Excellent | 50 (8.4%) | 130 (16.8%) | 180 (13.1%) |
| Socioeconomic status, mean IRSAD quintile (SD) | 3.7 (1.4) | 3.7 (1.4) | 3.7 (1.4) |
| Remoteness |  |  |  |
| Major cities | 438 (73.5%) | 589 (76.2%) | 1027 (75.0%) |
| Other | 158 (26.5%) | 184 (23.8%) | 342 (25.0%) |
| Adequate health literacy<sup>&</sup> | 505 (90.3%) | 665 (91.7%) | 1170 (91.1%) |
| eHealth literacy, mean (SD) | 4.2 (0.7) | 4.1 (0.7) | 4.2 (0.7) |
| Patient activation<sup>%</sup>, mean (SD) | 74.7 (13.2) | 75.0 (13.4) | 74.9 (13.3) |
| Cancelled/ postponed an appointment in the last 4 weeks because of COVID-19 | 147 (24.7%) | 125 (16.2%) | 272 (19.9%) |
| Felt needed to see a health professional in the last 4 weeks but chose not to | 115 (19.3%) | 104 (13.5%) | 219 (16.0%) |
| Needed to access a telehealth service in the last 4 weeks but could not | 12 (2.0%) | 7 (0.9%) | 19 (1.4%) |

<sup>^</sup> chronic health conditions include respiratory disease, asthma, COPD, hypertension, cancer, heart disease, stroke, diabetes. <sup>&</sup> based on Newest Vital Sign (NVS); data missing due to technical errors with Qualtrics for n=85 (6.2%) participants. <sup>%</sup> based on Consumer
Health Activation Index (CHAI); 0-79 low activation, 80-94 moderate activation, 95-100 high activation. eHealth literacy was measured on a 5-point Likert scale, a higher score reflects a higher level of eHealth literacy. Based on Index of Relative Socio-economic Advantage and Disadvantage (IRSAD) quintile (1 is most disadvantaged and 5 most advantaged).

#### Cancellation of in-person appointments

There were 272 respondents (19.9%) who cancelled or postponed an in-person appointment with a health professional. The reasons for cancelling appointments were due to: concerns about catching COVID-19 at a clinic or hospital (n=85, 31.3%), isolating due to COVID-19 symptoms or risk (n=31, 11.4%), concerns about travelling on public transport (n=21, 7.7%), feeling too busy (n=20, 7.4%), cost (n=9, 3.3%), and other reasons (n=106, 39%). Less frequent reasons for cancelling or postponing an in-person appointment included: border closures, elective surgery postponed, and the appointment seemed non-essential. A total of 219 respondents (16%) felt they needed to see a health professional in-person in the last 4 weeks but chose not to go, due to: concerns about catching COVID-19 at a clinic or hospital (n=72, 32.9%), feeling too busy (n=37, 16.9%), isolating due to COVID-19 symptoms or risk (n=18, 8.2%), concerns about travelling on public transport (n=13, 5.9%), other reasons (n=67, 30.6%). Less common reasons listed for choosing not to see a health professional included: only telehealth available, limited in-person appointment availability, and feeling too complicated.

#### Telehealth Experiences

The characteristics of the telehealth users experience are shown in Table 2. Of those who used telehealth, over half reported having more than one telehealth appointment (54.7%) most of which were delivered by telephone (72%). The majority of respondents (62%) rated their telehealth experience as “just as good” or “better” than a traditional in-person medical visit. On average, respondents perceived telehealth as moderately to very useful for medical appointments after the COVID-19 pandemic is over (M=3.67 out of 5, SD=1.1). Individuals who responded that their telehealth experience was *worse* than traditional in-person medical visits (34.4%) also rated the usefulness of telehealth after the COVID-19 emergency is over as significantly lower (2.86 vs 4.17, Mean difference=1.31, 95%CI: 1.14 to 1.47, t(572)=15.62, p<0.001) than those whose experience was just as good or better than an inperson visit.

**Table 2.** Characteristics of telehealth users experience (n=596). Data are presented as n (%) unless otherwise indicated.

| Variable |  | Summary Value<br>n (%) |
| --- | --- | --- |
| Number of telehealth appointments | One | 270 (45.3%) |
|  | Two | 157 (26.3%) |
|  | Three or more | 169 (28.4%) |
| Mode of telehealth delivery | Telephone | 427 (71.6%) |
|  | Videoconference | 84 (14.1%) |
|  | Both | 85 (14.3%) |
| Telehealth visit compared to traditional in-person medical visit | Better | 49 (8.2%) |
|  | Just as good | 320 (53.7%) |
|  | Worse | 205 (34.4%) |
|  | Unsure | 22 (3.7%) |

#### Factors associated with poorer telehealth experience

Multivariable analysis exploring factors associated with poorer telehealth experience compared to in-person appointments are displayed in Table 3. Male gender (p=0.007), having a history of both depression and anxiety (p=0.037), or a lower patient activation score (p=0.037) were associated with a poorer telehealth experience after controlling for all other variables in the model.

**Table 3.**
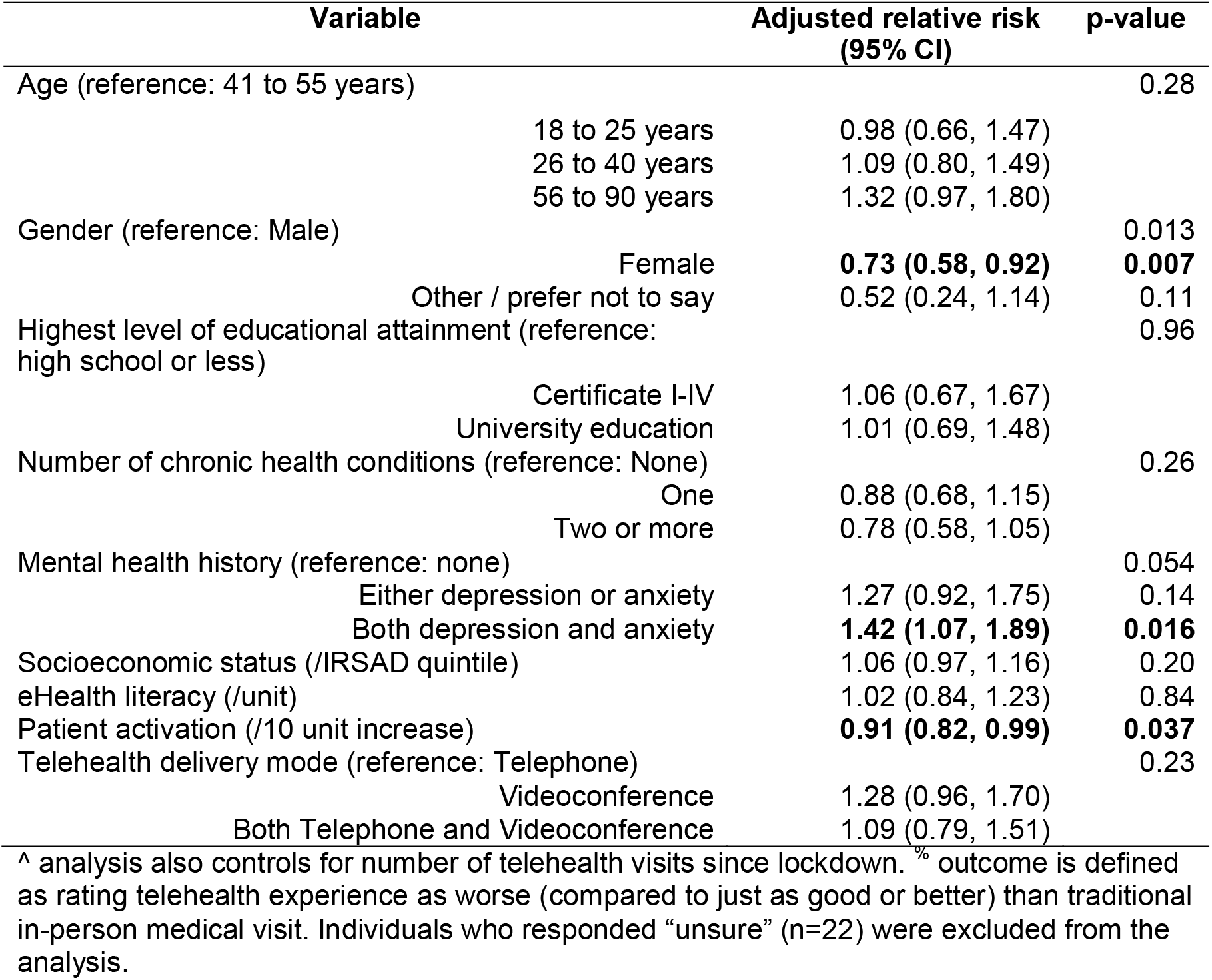
Multivariable^ analysis of factors associated with poorer^%^ telehealth experience compared to in-person appointment (n=574). Values displayed are relative risks (with 95% confidence intervals). Adjusted relative risks less than 1 indicate a reduced risk of reporting a poorer telehealth experience (relative to the reference group).

#### Reasons provided for why telehealth experiences were worse than traditional in-person visits

Six overarching themes (outlined in Table 4) emerged from the free-text data; 1) Communication is not as effective, 2) Limitations with technology, 3) Issues with obtaining prescriptions and pathology, 4) Reduced confidence in doctor, 5) Additional burden for complex care, and 6) Inability to be physically examined. Overall, the most recurrent theme was that communication was not as effective as traditional in-person visits, due to lack of visual cues, eye contact and body language.

**Table 4.** Reasons for telehealth visits being worse than traditional in-person medical visits, frequency of overarching thematic themes and subthemes with example quotes (n=221, 37%)*

| Code description | Example | N (%) |
| --- | --- | --- |
| **Communication is not as effective as face-to-face** |  |  |
| Lacks visual cues, eye contact, body language, visual feedback, prefers face-to-face | *"The subtle facial expressions eye contact and body language are not the same"* | 54 (24.4%) |
| Less personal, less natural/comfortable, more awkward | *"Difficult to establish rapport"* | 46 (20.8%) |
| Less effective/communication not as good, less helpful, harder/more difficult | *"Communication on the phone is less effective"* | 45 (20.4%) |
| Face-to-face interaction is needed for mental health appointments | *"I feel a big part of effective mental health care involves face-to-face conversation"* | 19 (8.6%) |
| More anxiety provoking for some | *"Phone call and videos make me extremely anxious"* | 5 (2.3%) |
| Less privacy | *"Lack of privacy"* | 3 (1.4%) |
| **The inability to be physically examined** |  |  |
| Physical exam is not possible | *"Could not have a physical exam done"* | 60 (27.1%) |
| Tests could not be performed | *"Blood pressure not taken"* | 17 (7.7%) |
| **Limitations with technology** |  |  |
| Technology issues including poor connection, bad reception, poor audio quality zoom dropped out | *"Due to audio quality I was not able to get names of chemotherapy drugs correctly - so when I tried to look up info later I couldn't until I was able to get info from Breast care nurse so this added to days of anxiety due to lack of info over weekend and when that staff member on leave."* | 20 (9.0%) |
| Poor quality connection led to poor quality conversation | *"Harder to communicate due to tech difficulties, lag issues"* | 6 (2.7%) |
| **Issues with obtaining prescriptions and pathology** |  |  |
| Harder to obtain prescriptions | *"I had to wait for scripts to be emailed to the pharmacy, then one was missing, which I could have seen at the time had I received them in person."* | 10 (4.5%) |
| Increased wait time/access delayed | *"If you need a script or referral, you have to make a separate trip to go get the paper"* | 7 (3.2%) |
| Unable to access pathology | *"Getting blood tests has become more difficult."* | 2 (0.9%) |
| **Reduced confidence in doctor/ health professional** |  |  |
| Not as comprehensive or thorough | *"Not as comprehensive and thorough"* | 25 (11.3%) |
| Time Pressure | *"Felt rushed"* | 18 (8.1%) |
| Lack of confidence in assessment, diagnosis | *"Less trust that the diagnosis is accurate"* | 11 (5.0%) |
| **Additional Burden for Complex Care** |  |  |
| Face-to-face required for complex issues | *"I had to go in for a face-face consults because the medical issues could not be diagnosed"* | 15 (6.8%) |
| Complex issues delayed | *"More complex issues have been delayed until we can do face-to-face"* | 5 (2.3%) |
| Added burden of having two consults | *"In both instances after having the Telehealth calls, I had to go in for a face to face consults because the issues could not be diagnosed over the phone"* | 4 (1.8%) |

\*Full text could have more than one theme applied

#### Barriers to Telehealth

Less than 2% of respondents reported not being able to access a telehealth service (n=19, 1.4%). The reported barriers were: telehealth was not available from their GP or health professional (n=4), they did not have internet (n=2), the appointment was not available when required (n=8) and that using telehealth felt too complicated (n=5).

## Discussion

Our findings showed that more than half of the respondents rated their telehealth experience as just as good or better than traditional in-person medical care. This is encouraging considering that community transmission of COVID-19 across Australia may continue to persist for some time. Of those who used telehealth, most anticipated that telehealth would be at least somewhat useful after the COVID-19 pandemic is over, suggesting telehealth may be a viable long-term option for healthcare delivery. Our findings are consistent with a survey reporting that 85% of older Australians found their telehealth experience to be similar or better than face-to-face consults (15). We found that telehealth modality (telephone vs video) was not associated with a poorer telehealth experience compared to in-person appointments, consistent with other studies showing telephone and videoconferencing to be comparable in terms of patient satisfaction (16). Perhaps unsurprisingly, individuals who rated their telehealth experiences as worse were less likely to perceive that telehealth would be useful beyond the COVID-19 pandemic. Those who rated their telehealth experience as worse than traditional-in person visits were more likely to be male, have lower patient activation (individuals’ willingness to take on the role of managing their health and healthcare), or have a history of both depression and anxiety. This last observation is also supported by the content analysis, which highlighted that traditional in-person visits were frequently preferred over telehealth visits for mental health appointments.

Our findings suggest that telehealth is less effective for mental health services. This is of concern as mental health problems such as depression and anxiety were at least twice as prevalent in the first month of the COVID-19 pandemic compared to before the pandemic in Australia (17), a problem that is only expected to grow (18). Our findings are similar to previous research on telehealth and mental health (19), which is concerning as negative experiences of telehealth may result in no mental health care for patients at all if face-to-face services are unavailable.

The most common theme for why respondents perceived telehealth to be worse than inperson medical care was less effective communication. This issue could be addressed by encouraging the use of established strategies to improve communication between health professionals and patients. For example, the teach-back method, also known as “show me” or “closing the loop”, has shown to increase people’s understanding of health information by asking the patient to repeat back health information in their own words (20). In addition, providing a written lay summary of the visit via a patient letter or patient portal may improve the telehealth experience for patients. Patients have reported improved patient-provider communication as a result of using a patient portal (21). Other approaches to address this issue may include online education or mobile applications, both of which have been used to enhance patient understanding of content and improve healthcare services (22, 23). It should be noted though that the Australian government is currently fast-tracking electronic prescribing which may improve communication between GP, patients and the pharmacist (24). This process allows easy electronic sharing of prescriptions, eliminating issues related to the challenge of obtaining prescriptions from telehealth appointments.

Additional issues identified in our study were that physical examination was not possible; people were less confident in their doctor/healthcare professional during telehealth; and additional burden was experienced for complex health conditions. These issues could be addressed by setting clear expectations for telehealth when scheduling appointments and expectations about which types of appointments are suitable for telehealth. For example, when appointments are scheduled, patients should be notified that depending on the complexity of their medical appointment, additional in-person consults may be required, or that telehealth may not be appropriate for mental health issues. In addition, video conferencing could be offered as it may help with more reliable visual assessment and greater diagnostic accuracy (16). Overall, in order to improve patients’ experiences with telehealth, strategies should be implemented to ensure that patients are aware of what to expect from telehealth appointments.

Importantly, our study identified that ~20% of respondents had cancelled or postponed an inperson health appointment, with the main reason for cancelling due to concerns about catching COVID-19 at health or hospital clinics. Our study identified a higher proportion of people who cancelled or postponed a health appointment than the survey conducted by the ABS (11%) (4). Similarly, a study in Israel of 151 women with breast cancer found that 31% of people cancelled a health appointment, with the most common reason being due to fear of contacting COVID-19 (25). This is worrying as continuing outbreaks may deter patients from accessing essential in-person medical care for some time. Therefore, our results suggest that telehealth services should continue to be offered while community transmission of COVID-19 persists. Future studies should investigate whether patients who are cancelling or postponing health appointments are seeking telehealth services and monitor the long-term impact of health service utilization on health outcomes

Whilst the study sample was large and diverse, it is not statistically representative of the Australian population, in particular including a higher proportion of females, a higher level of education, and potentially higher levels of digital literacy. In addition, our survey did not collect any information on the type of telehealth services that participants attended (e.g. allied health, specialist). Future surveys should investigate how people’s experience with telehealth compares to traditional in-person visits depending on the health service type (e.g. GP, Specialist, Allied Health) and determine the impact of different health service modalities and utilization on health outcomes.

## Conclusions

Overall, we found that telehealth was reported to be comparable to traditional in-person visits by most of our participants. We identified the most common reasons for a poor experience of telehealth and provided strategies which could help to improve the experience of telehealth users. Universal telehealth should continue to be offered as a mode of healthcare delivery while the pandemic continues and may be worthwhile beyond the pandemic.

## Data Availability

Data is available upon reasonable request.

## Acknowledgements

We would like to acknowledge the members of the Australian public who participated in this survey.

No funder had any role in the study.

There are no conflicts of interest.

All authors had full access to all of the data including statistical reports and tables.

